# A Bayesian network model for personalised COVID19 risk assessment and contact tracing

**DOI:** 10.1101/2020.07.15.20154286

**Authors:** Norman E Fenton, Scott McLachlan, Peter Lucas, Kudakwashe Dube, Graham A Hitman, Magda Osman, Evangelia Kyrimi, Martin Neil

## Abstract

Concerns about the practicality and effectiveness of using Contact Tracing Apps (CTA) to reduce the spread of COVID19 have been well documented and, in the UK, led to the abandonment of the NHS CTA shortly after its release in May 2020. We present a causal probabilistic model (a Bayesian network) that provides the basis for a practical CTA solution that addresses some of the concerns and which has the advantage of minimal infringement of privacy. Users of the model can provide as much or little personal information as they wish about relevant risk factors, symptoms, and recent social interactions. The model then provides them feedback about the likelihood of the presence of asymptotic, mild or severe COVID19 (past, present and projected). When the model is embedded in a smartphone app, it can be used to detect new outbreaks in a monitored population and identify outbreak locations as early as possible. For this purpose, the only data needed to be centrally collected is the probability the user has COVID19 and the GPS location.

## 1. Introduction

Central to many governmental measures to contain the current COVID19 infection and reduce the impact of potential subsequent waves, are the use of manual and smartphone-based contact tracing methods, referred to as Contact Tracing Apps (CTA). This paper is part of a set of studies conducted by a multidisciplinary team of researchers that suggest that any standalone contact tracing approach, manual or automated, is unlikely to contain, a high-prevalence highly contagious disease like COVID19 (McLachlan et al, 2020b; McLachlan et al 2020c).

The first of these studies (Scott McLachlan, Lucas, Dube, Hitman, et al. 2020) conducted a systematic review of recent literature proposing CTA solutions for COVID19, highlighting issues relating to privacy and confidentiality and identifying the *sweet-spot* for the uptake of CTA at which it becomes clinically useful. We showed that the best-case scenario for uptake requires between 90% and 95% of the entire population for infection containment. This does not factor in any loss due to people dropping out or device incompatibility or the fact that only 79% of the population own a smartphone, with less than 40% in the over-65 age group. Hence, the best-case scenario is beyond that which could conceivably be achieved, demonstrating that CTA on its own is not adequate as the sole containment tool in a pandemic. The framework of our proposed solution was described in (Scott McLachlan, Lucas, Dube, McLachlan, et al. 2020). We argued that, in addition to privacy issues, the UK’s adopted CTA solution (NHSX) (Servick 2020) only acts retrospectively, advising the user they were previously in close contact with another infected person. This advice often comes only after they have already begun asymptomatically shedding the disease (shortly after our review the UK Government announced that it was no longer supporting NHSX and had started to work on an alternative solution with Apple and Google^1^). In contrast, the solution we proposed integrates any retrospective CTA with symptom tracking and a causal probabilistic model (a Bayesian network) that computes the probability of COVID19 infection based on information provided by the individual and publicly available data. Our solution does not compromise privacy because information provided by the user is not stored on any central server. We believe this solution could work when combined with an effective and transparent communication strategy. In (Osman et al. 2020) we addressed the challenges of both technical and behavioural issues in adopting CTA that policy makers need to consider to successfully deal with the possibility of a second or subsequent wave.

In this paper we describe the details of the proposed Bayesian network model and its deployment first introduced in (Scott McLachlan, Lucas, Dube, McLachlan, et al. 2020). The model focuses on computing the probability that a person has asymptomatic, mild or severe COVID19 (past, present and projected). The model and its assumptions are described in Section 2. In Section 3 we provide several scenarios demonstrating how the model makes diagnoses and predictions and provides personalised decision-support for users. For instance, it provides personal decision-making and answers questions, such as ‘*whether or not I have the disease?*’, ‘*whether or not I should go out?’*, and ‘*Whether or not it is safe to be in contact with others?’* (Osman et al. 2020). We also demonstrate how to perform sensitivity analysis to determine which variables in the model have most impact on any particular outcome variable. Limitations of the model are discussed in Section 4. We propose that this model takes the form of smartphone app based on the architectural framework that we describe in Section 5 (this architecture is already in operation as part of an EPSRC project^2^). The app can be used to detect where new outbreaks occur as early as possible. For this purpose, the only data needed to be stored centrally is the probability the user has COVID19 and the GPS location. In particular, none of the personal factors entered into the model by the users are stored centrally. Hence – unlike other proposed solutions – there is less possibility of compromising users’ privacy.

## 2. Bayesian network based COVID19 symptom tracking and prediction^3^

A Bayesian network (BN) (Cowell et al. 1999; Pearl 1988; Fenton and Neil 2018) is a graphical model, consisting of nodes and arcs, as shown in Figure 1 (which is a very simplified fragment of the main COVID19 model we will present). The nodes represent variables which may or may not be directly observable (the convention we use here is that those coloured white are generally not observable). Nodes may be discrete or continuous (for simplicity we consider only discrete nodes here) and the probability values displayed represent the probabilities associated with the state values of that node.

**Figure 1.**
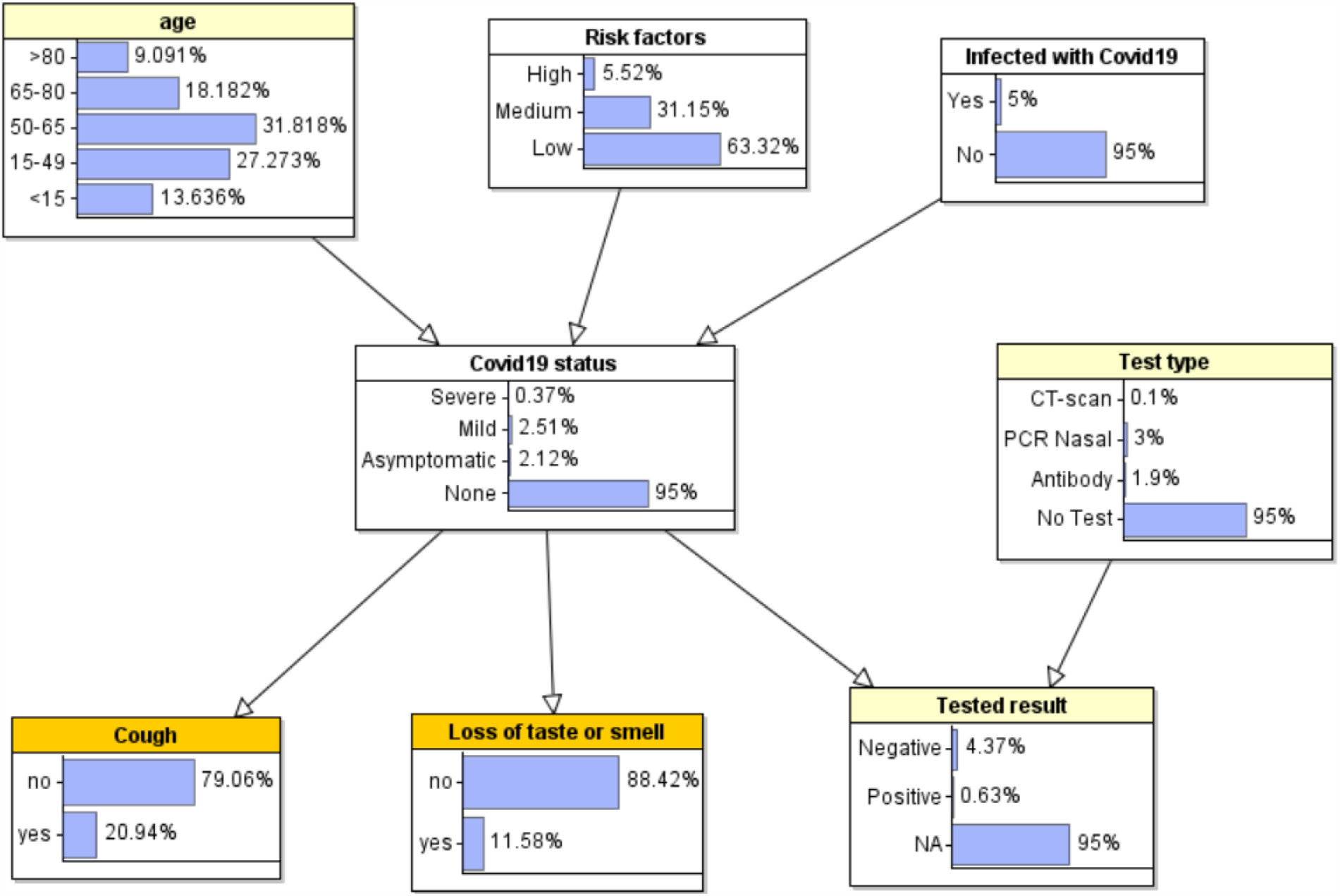
A Bayesian network (this is a very simplified fragment of the main COVID19 Bayesian network model structure). The probabilities shown represent prior marginal probabilities when no observations are entered

There is an arc between two nodes if the corresponding variables are causally or statistically linked. The strength of the link, as well as the uncertainty associated with these, is captured in probability tables such as those shown in Table 1 and Table 2. The values (formally called ‘parameters’) in these tables are either learned from data or knowledge - or a combination of both. Every node which has at least one parent will have an associated probability table that is conditioned on its parent node(s). For example, the probability table for the node “Tested result” is shown in Table 1 and is based on a combination of data from empirical studies (Carver and Jones 2020; Ai et al. 2020; Yang et al. 2020; Chua et al. 2020) and knowledge such as the obvious fact that ‘No Test’ will not produce a result. The table for the node ‘Cough’ is shown in Table 2 and is based on a combination of data from (Michelen, Jones, and Stavropoulou 2020) and knowledge.

**Table 1.** Conditional probability table for node “Tested Result”

| Test type | CT-scan |  |  |  | PCR Nasal |  |  |  | Antibody |  |  |  | No Test |  |  |  |
| --- | --- | --- | --- | --- | --- | --- | --- | --- | --- | --- | --- | --- | --- | --- | --- | --- |
| Covid19 status | Severe | Mild | Asymp... | None | Severe | Mild | Asymp... | None | Severe | Mild | Asymp... | None | Severe | Mild | Asymp... | None |
| Negative | 0.01 | 0.02 | 0.02 | 0.99 | 0.1 | 0.3 | 0.46 | 0.9 | 0.3 | 0.3 | 0.3 | 0.9 | 0.0 | 0.0 | 0.0 | 0.0 |
| Positive | 0.99 | 0.98 | 0.98 | 0.01 | 0.9 | 0.7 | 0.54 | 0.1 | 0.7 | 0.7 | 0.7 | 0.1 | 0.0 | 0.0 | 0.0 | 0.0 |
| NA | 0.0 | 0.0 | 0.0 | 0.0 | 0.0 | 0.0 | 0.0 | 0.0 | 0.0 | 0.0 | 0.0 | 0.0 | 1.0 | 1.0 | 1.0 | 1.0 |

**Table 2.** Conditional probability table for node “Cough”

| Covid19 status | Severe | Mild | Asymptomatic | None |
| --- | --- | --- | --- | --- |
| no | 0.29 | 0.343 | 0.99 | 0.8 |
| yes | 0.71 | 0.657 | 0.01 | 0.2 |

For nodes that have no parents (such as the node “Age”) the associated probability table is simply based on population statistics. For example, from national UK statistics 18.1% of the population is aged between 65 and 80 and hence the probability value associated with the state “65-80” is 18.1%. Such probabilities are called ‘prior’ probabilities. For nodes with parents, such as the node ‘cough’ or the node ‘COVID19 status’, the probability values shown in Figure 1 are called ‘marginal prior probabilities’; they are calculated based on the values of their parent nodes and their conditional probability table. Hence, the fact that the prior marginal probability for ‘COVID19 status’ mild being 2.51% tells us that, based on the current population distribution of age, risk factors and disease infection, 2.51% of the population currently has mild COVID19.

So, while the initial probability values in a BN model represent what is known about the population as a whole, the real purpose of such a model is to ‘personalize’ it by providing it with information specific to an individual. For example, if we know that a specific individual is aged between 50-65 then we can enter this information into the model – we call this ‘entering an observation’. When observations are entered into the model for specific variables, all of the probabilities for the, as yet unobserved variables, are updated using an AI algorithm called Bayesian inference. For example, Figure 2 shows the updated probabilities when a user enters the following observations about themselves (or, say, a family member): a) they are aged 50-65; b) they have symptoms of a cough and loss of taste and smell. So, whereas the ‘prior’ probability for COVID19 status being ‘mild’ was 2.51% (Figure 1) the revised probability – which we call the ‘posterior probability’ - has now increased to 42.2%.

**Figure 2.**
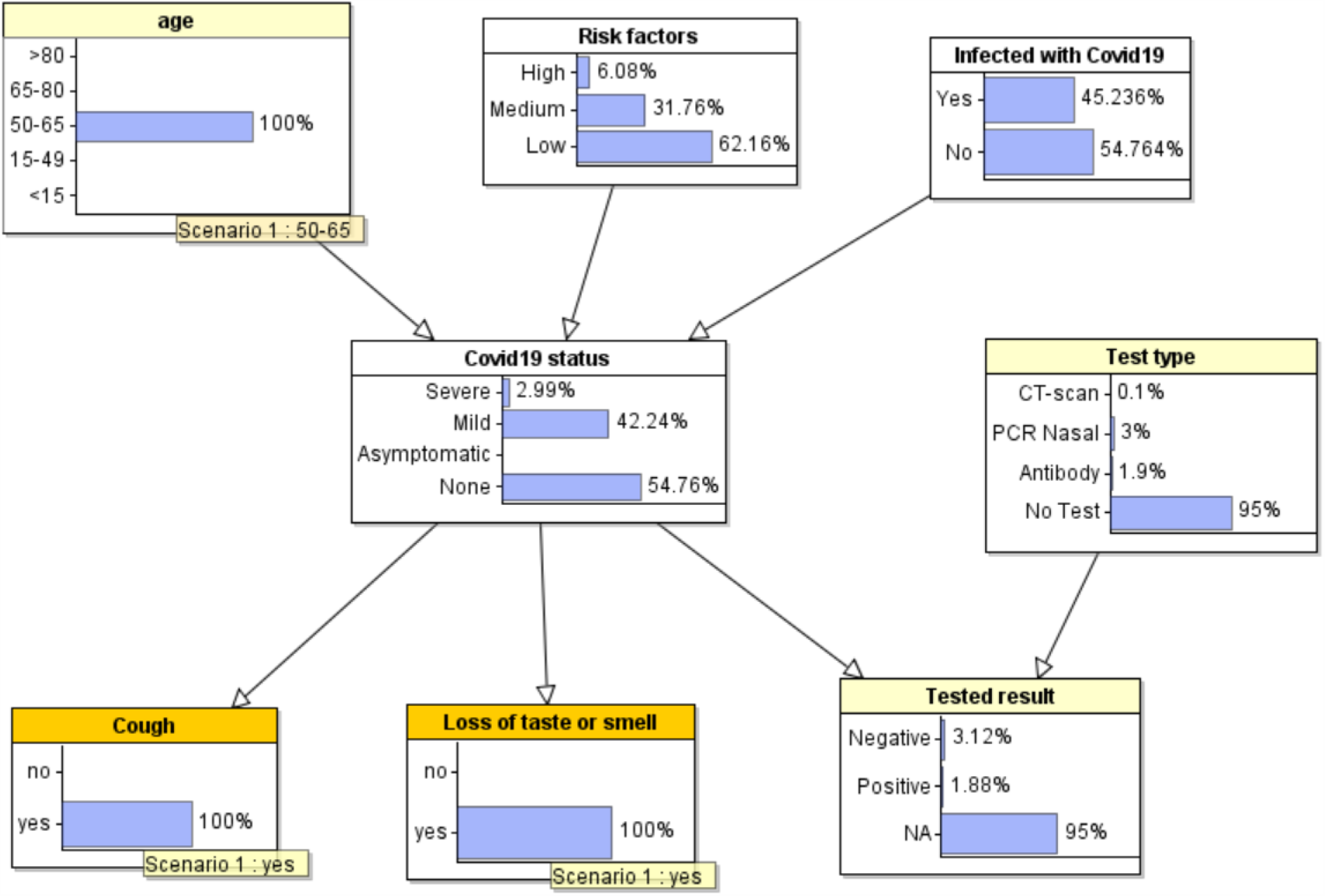
Updated probabilities when observations are entered

It is important to note that we call models such as this *causal* BNs to distinguish them from BN models whose structure (and probability parameters) are learnt entirely from data - the limitations of learning BN models solely from data for problems such as this are well known (Constantinou et al. 2016). The model structure here is defined by basic knowledge of causation that confirm to a set of well-defined structural idioms (Kyrimi et al. 2020; Neil, Fenton, and Nielson 2000) that themselves represent reusable patterns of causal inference. For example, symptoms such as loss of taste or smell in this model (Centers for Disease Control and Prevention 2020) do not *cause* a particular disease status, but are caused by it. Hence, the arc is directed from disease status to symptoms and not vice versa. Conversely, physiological risk factors (such as age or existing medical conditions) may increase or decrease the probability of a disease status but are not caused by the disease. Hence, the arcs are directed from risk factors to disease status.

It is also important to note that the model we present below is quite different to the kind of BN models we used in our recent publications (Neil et al. 2020; Fenton et al. 2020) about COVID19. Those models were aimed at learning, from data, the ‘unknown’ infection and death rates over entire populations, taking account of uncertain information such as accuracy and bias in testing and death reporting. So, nodes^4^ in those models represented the (unknown) infection and death rates and test accuracy results. In contrast, the model described here does not allow us to properly incorporate uncertainty about population infection rates and test accuracy. Instead, we rely on ‘point’ probability values entered into relevant probability tables. For example, the probability table for the node ‘Tested result’ shown in Table 1 contains only point values (as opposed to full probability distributions) for the various false positive and false negative rates associated with the different types of tests. These point values represent the average over the various reported values and are thus a simplification. However, because this model is concerned with predictions about an individual rather than a population, these simplifications are reasonable.

The structure of the full COVID19 model is shown in Figure 3. The node states and prior marginal probability values are shown in Figure 4; that is, the state of the model when no observations are entered. This reflects a current random individual in the general ‘population’.

**Figure 3.**
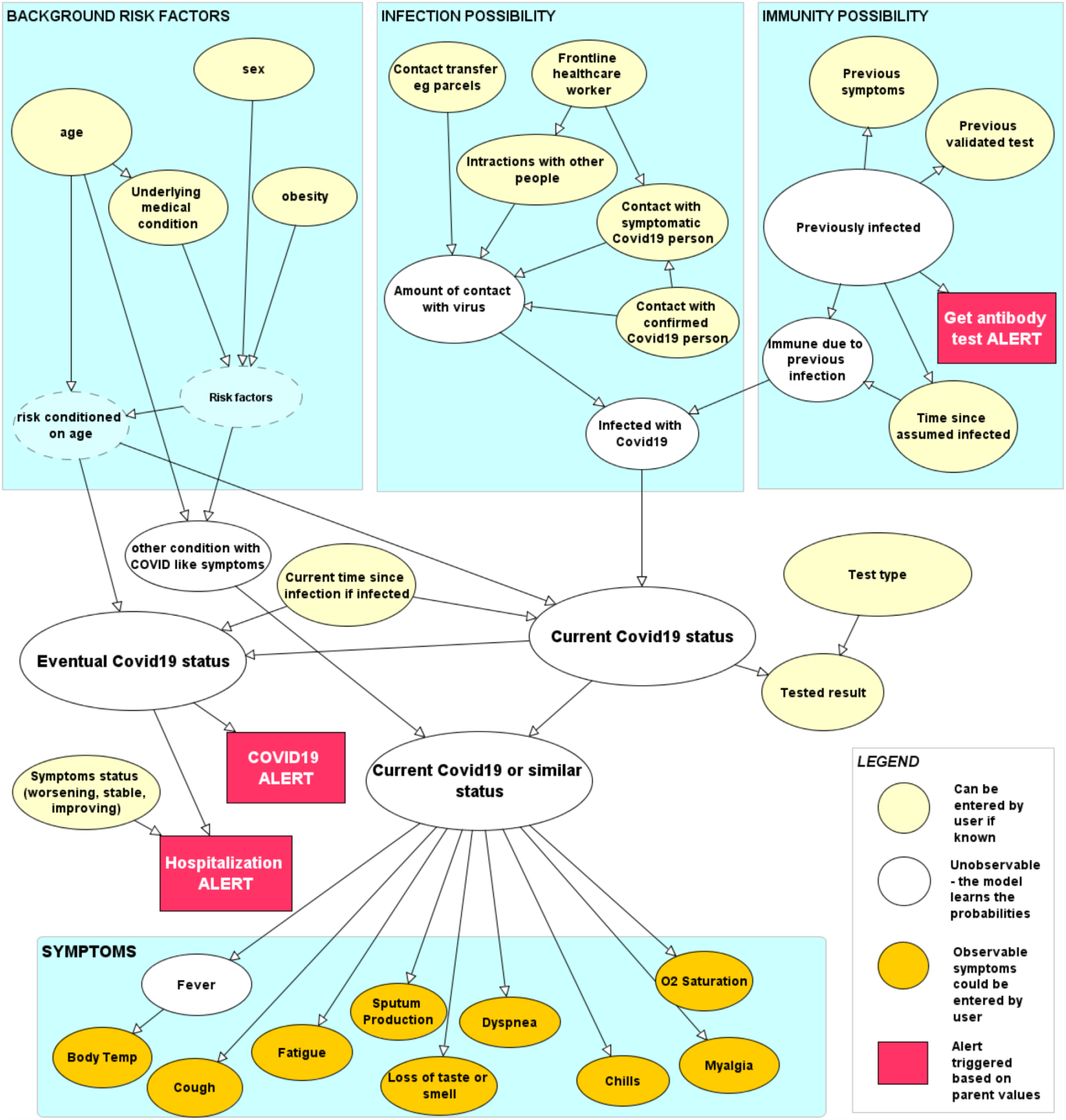
Structure of the COVID19 model

**Figure 4.**
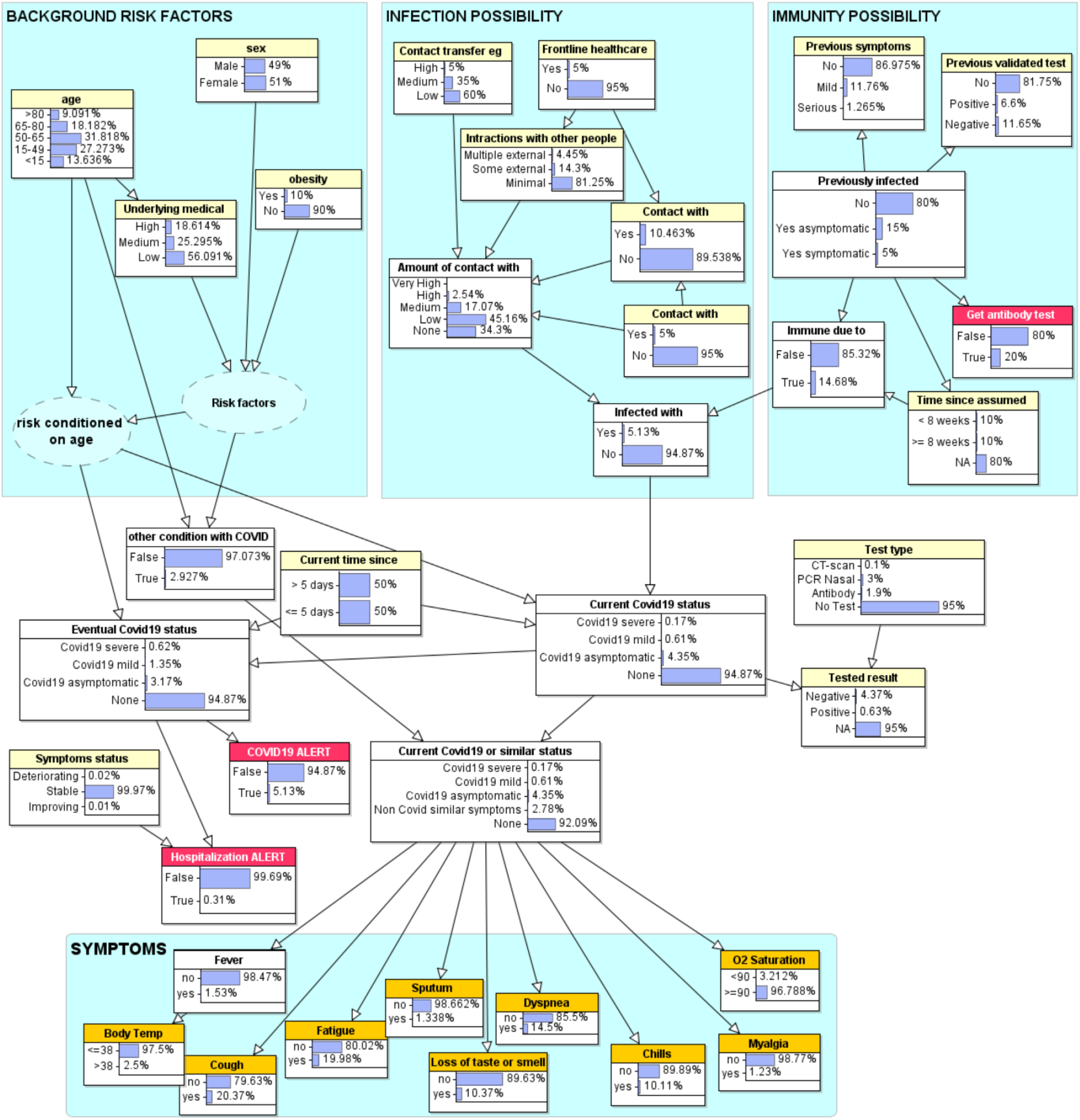
Model with prior marginal probabilities

The assumptions and rationale for the model are as follows:

- A person can only become infected if they have encountered the virus, and the amount of contact with the virus determines the probability of actual infection. Hence, we have a part of the model (“Infection”) that focuses on the amount of contact a person has recently had with the virus (where amount is assessed on a simple ranked scale from none to very high). For someone who knows they have been in direct contact with a confirmed COVID19 person the amount of contact with the virus will likely be high. However, most people will be unaware of whether or not people they have been in contact with were infected. They might know if people they were with had symptoms; they should also be able to say how much general interaction they have had with people outside their household (such as through work, and attending parties and larger gatherings) and whether there may have been contact transfer of the virus to them from, for example, receiving many recent parcel deliveries. Any information about these will update the probability of infection.
- A person previously known to have had COVID19 may have immunity from contracting it again. Hence, we have a part of the model (“Immunity”) that focuses on evidence to support the notion that a person was previously infected. Typically, people assume that they were previously infected if they previously had severe symptoms and/or had a validated test that was positive. It is those observations that a user can enter into the model (rather than a direct observation that they were infected); the model captures uncertainty about the accuracy of both types of evidence. The model also incorporates empirical information about the decreasing impact on immunity given time since infection (Long et al. 2020). For simplicity we differentiate only between less than 8 weeks and at least 8 weeks.
- The listed background risk factors (age, sex, obesity and underlying medical conditions) have no impact on whether or not a person becomes infected. However, if a person is infected then these risk factors do impact on whether or not the infection will be asymptomatic, mild or serious.
- If a person becomes infected with COVID19 then in the first 5 days they are generally asymptomatic (Beeching, Fletcher, and Beadsworth 2020). Some infected will never get any symptoms (i.e. they remain asymptomatic) (Heneghan, Brassey, and Jefferson 2020), whereas for others the disease will eventually be classified as either mild or severe. So, we have separate nodes distinguishing between the current COVID19 status and eventual worst case COVID19 status (Mizumoto et al. 2020; Day 2020; Bengali 2020).
- In general, we will never observe directly that we have or have not been infected with the virus. So, we would never enter a direct observation of yes or no into the node ‘infected with COVID19’ (nor into the node ‘previously infected’). However, if the probability of yes is zero then, irrespective of any other observations in the model the COVID19 status will be ‘none’ (even in the presence of the worst risk factors and full set of symptoms).
- There are other medical conditions (notably Flu and COPD) that have several similar symptoms to COVID19. Hence, the parent of the symptom nodes includes a state “Non Covid similar symptoms” to incorporate all such conditions. It is possible to have both COVID19 and a different condition.
- There are three major types of test (as shown in Figure 4) with different degrees of accuracy (characterised by the false positive and false negative rates defined in the probability table of “Tested result”). Although most people have not been (and will not be) tested^5^, if they have been then it is assumed they will know the result (and normally also the type of test) and enter this as an observation. The test accuracy data is based on (Carver and Jones 2020; Ai et al. 2020; Yang et al. 2020; Chua et al. 2020).
- Apart from the various factors relevant to the infection possibility (i.e. those associated with contact with infected people or objects) the only risk factors explicitly considered are age, sex, underlying medical condition and obesity. An extended version of the model that includes many other risk factors – such as diabetes and those associated with BAME communities and/or deprivation (Apea et al. 2020) - based on recent empirical data is described in (Prodhan and Fenton 2020). However, the dangers of including many claimed risk factors such as those in (Collaborative et al. 2020) are highlighted in (Griffith et al. 2020; Fenton 2020).
- The set of symptoms included are those occurring in at least 25% of patients with either mild or severe COVID19. The probability values are based on the statistics provided in (Huang et al. 2020). An extended version of the model incorporates many more l symptoms such as nausea, vomiting and diarrhoea, congestion and runny based on recent empirical data (Butcher and Fenton 2020).
- The two nodes with dotted lines (‘risk factors’ and ‘risk conditioned on age’) are introduced into the model solely for the benefit of simplifying parameter estimation. They are dotted because they are considered to be ‘hidden nodes’^6^ of no interest to users of the model.
- A threshold (such as 60%) is set for the nodes “COVID19 alert” and “hospitalisation alert”. If the former is triggered it will provide advice about submitting for a PCR nasal test as soon as possible, self-isolation and social distancing. The latter depends on the probability of severe (as opposed to mild or asymptomatic COVID19) and whether or not the symptoms are deteriorating. The node ‘Symptoms status’, which is a parent of ‘Hospitalization alert’ is assumed to consider factors such as: trouble breathing, persistent pain or pressure in the chest, new confusion, inability to wake or stay awake, bluish lips or face (Centers for Disease Control and Prevention 2020). There is also a “Get antibody test” alert for those who have a high probability of being previously infected.

## 3. Scenarios running the model

Figure 5 shows a scenario in which a 70-year-old person with serious underlying medical conditions (who is not a frontline healthcare worker and has not previously had a validated test or symptoms) has in the past 5 days had multiple recent interactions with other people including one with COVID19 symptoms.

**Figure 5.**
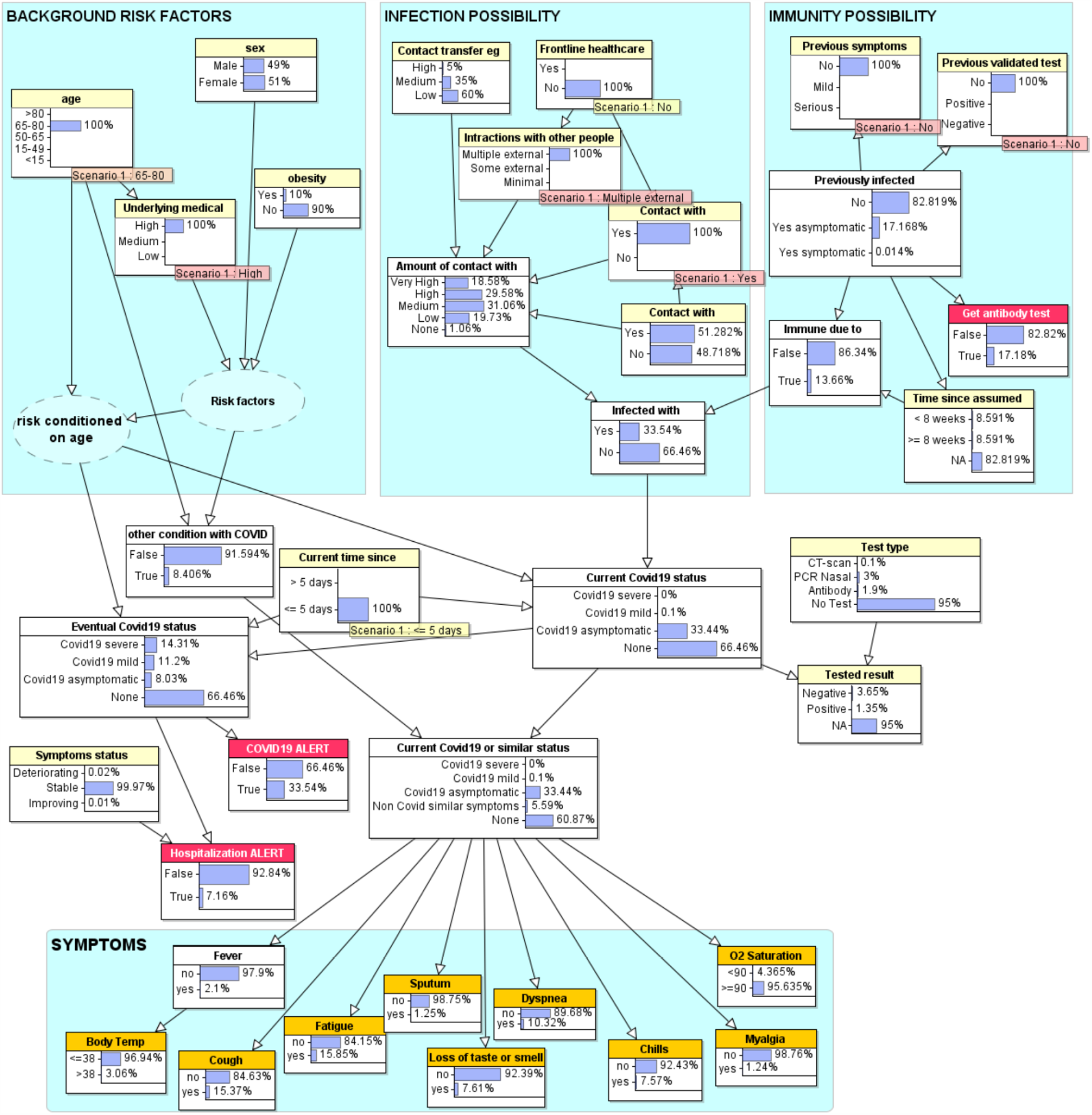
Scenario in which a person who is not a healthcare worker but has had multiple external interactions with others including one with COVID19 symptoms.

The model calculates there is a 33.5% probability this person is now infected. As the interactions were less than 5 days ago there are no currently reported symptoms. However, because of this person’s age and underlying medical conditions, the model predicts that (although currently asymptomatic) there is a 14% chance this person will eventually have severe COVID19 and therefore could be advised to self-isolate until they receive the test result

The particular observations for age and underlying medical conditions do not, as we made clear in our assumptions, change the probability of being infected, but it does change the probability of the eventual seriousness of infection if infected. Table 3 shows some examples of the results running the model with different age/medical condition combinations.

**Table 3.** Results of running the scenario with different age, underlying medical condition combinations

| **Age** | **underlying medical conditions** | **Eventual COVID19 status** |  |  |
| --- | --- | --- | --- | --- |
|  |  | **severe** | **mild** | **asymptomatic** |
| >80 | High | 20% | 9% | 5% |
| 65-80 | Medium | 14% | 11% | 8% |
| 15-49 | Medium | 2% | 8% | 24% |
| 15-49 | Low | 1% | 4% | 29% |
| <15 | Low | 0.15% | 0.65% | 33% |

If, in this scenario, we conduct a PCR nasal test and get a positive result the probability of COVID19 rises to 73%. If it was a CT-Scan positive test then the probability rises to over 98%. However, even in the latter case for a person aged between 50-65 with no underlying medical conditions, the model predicts only a 6% chance that the infection will be severe (27% mild, 65% asymptomatic) and therefore the appropriate medical advice can be given by a health professional or advice line.

Next we consider a scenario, shown in Figure 6, where a person aged between 50-65 with no underlying medical conditions reports symptoms: cough, fatigue, and dyspnoea. Note that, because this person has provided no information about recent contact with others and because the symptoms reported are also common generally (especially for those with colds or flu), the model calculates only a 6% probability of COVID19 (and 2.3% probability of a condition like flu).

**Figure 6.**
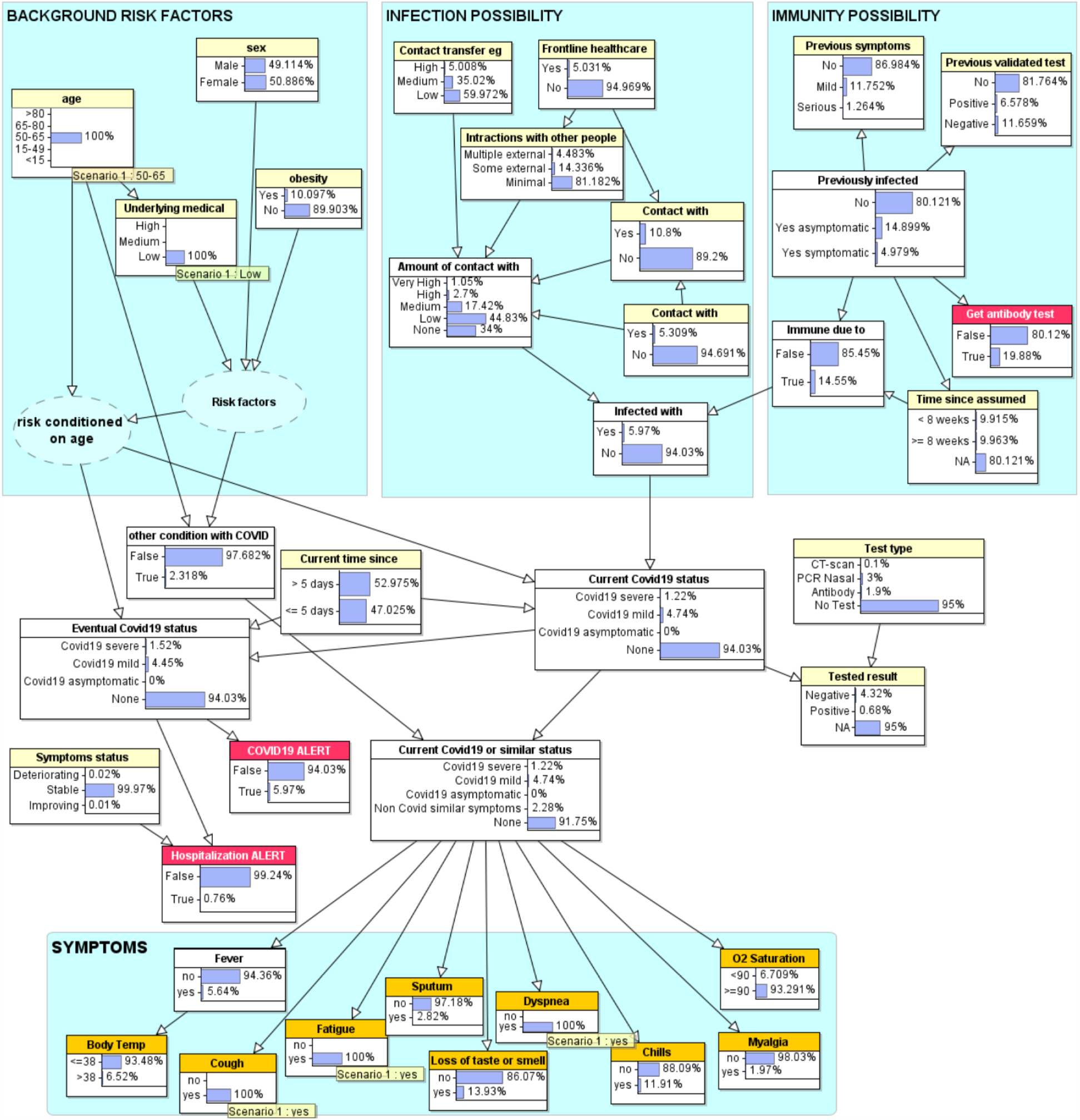
Person aged between 50-65 with no underlying medical conditions reports symptoms: cough, fatigue, dyspnoea

However, providing further symptomatic information can make a big difference. For, example if this person explicitly reports no loss of taste/smell (as opposed to not reporting anything for this symptom) the probability of COVID19 drops to 2%, whereas it jumps to 31% if there is loss of taste/smell. If in the latter case we further know that this person is a frontline healthcare worker, the probability increases to 44%. If this person has a PCR nasal test and gets a negative test result, there is still a 18% chance this person has COVID19 (the probability of a different flu-like condition is 4%) and therefore the advice might be to self-isolate for 7-14 days to further monitor symptoms and to have a repeat test prior to coming back to work.

In Figure 7 we see the result of running the model for a person about whom we have no observation other than that they have extensive symptoms, which are deteriorating.

**Figure 7.**
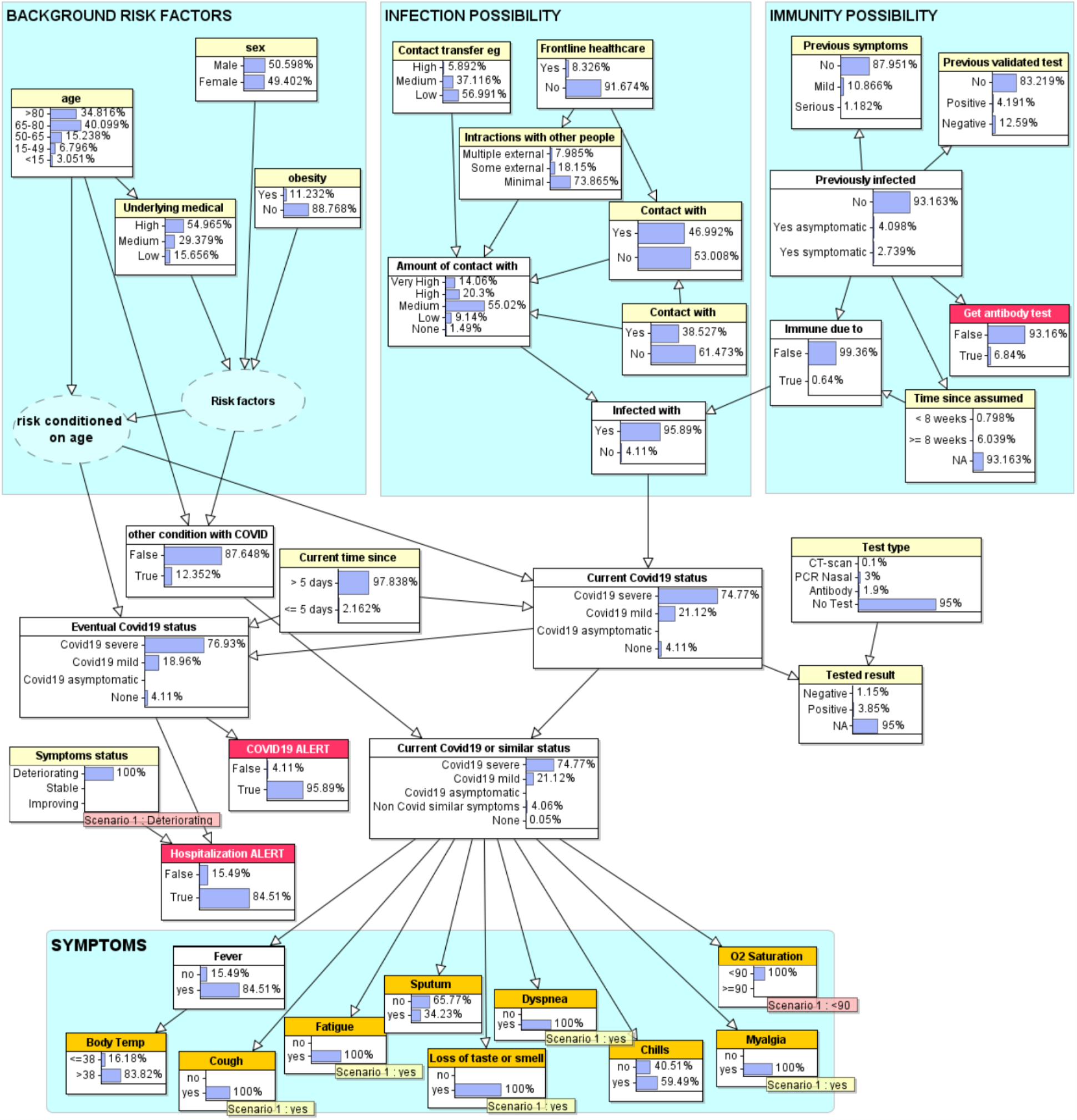
Person about whom we know nothing other than extensive symptoms

In this case the Bayesian algorithm’s ‘backward inference’ not only produces a very high eventual probability (77%) of severe COVID19 (19% mild), but also infers this person is very likely to be at least 65 with underlying medical conditions. As such, note that the hospitalization alert would be enabled if, say, the threshold was set at 60%. However, if we discover this person is, say only 45 with no underlying medical conditions, then although the probability of COVID19 is unchanged the probability that the worst-case scenario for this person is severe COVID19 drops from 77% to 28%; even with deteriorating symptoms, the hospitalization alert probability is below the 60% threshold.

Figure 8 shows the results of running the sensitivity analysis tool on the model in AgenaRisk for the ‘eventual COVID19 status’ (which is what we call the ‘target node’ for the sensitivity analysis) being severe. This assumes no observations in the model.

**Figure 8.**
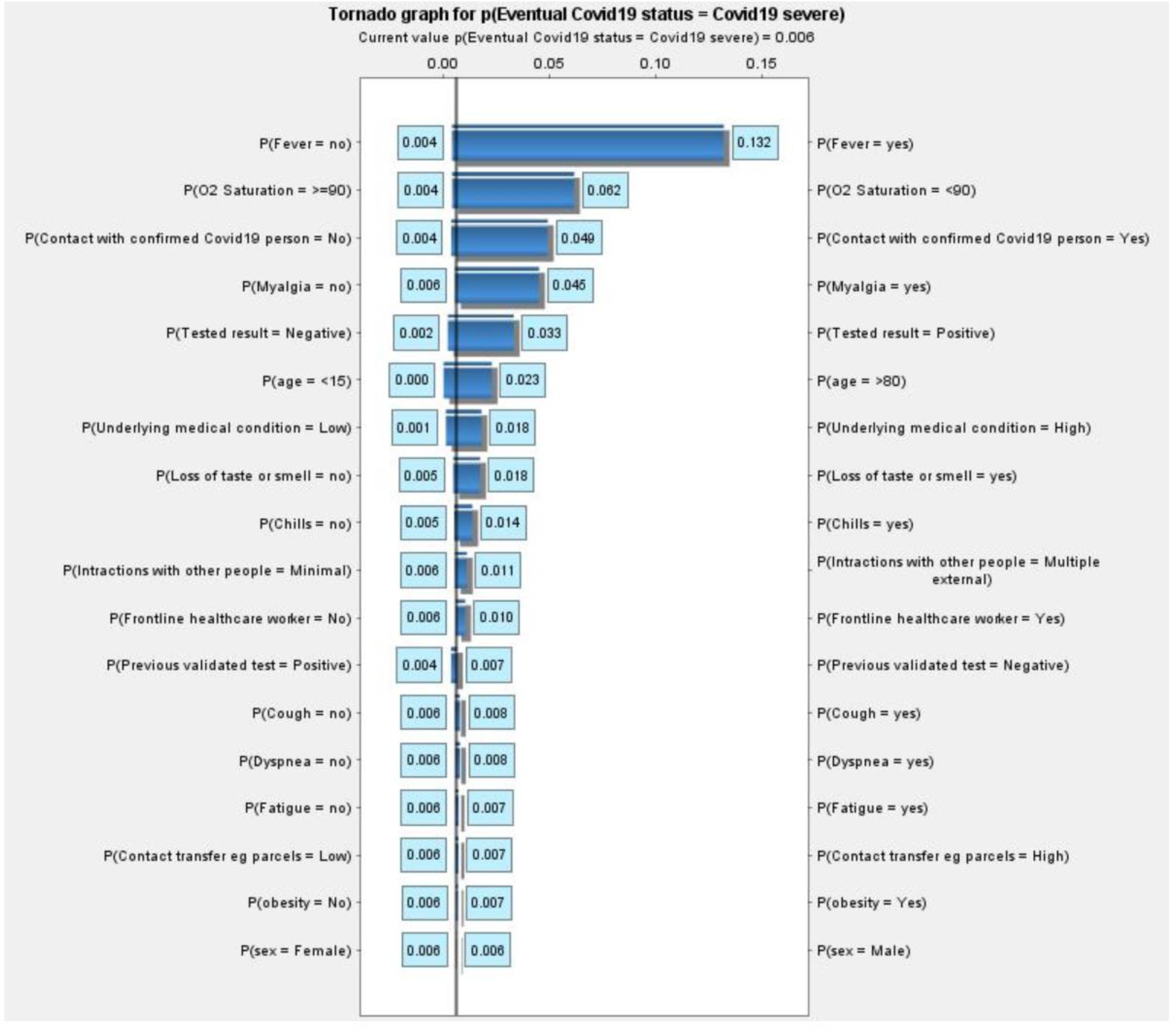
Sensitivity analysis for “eventual COVID19 status = severe” shown as tornado graph

It is possible to run many different types of sensitivity analysis. For example, once some observations are entered for a specific patient, we could run the sensitivity analysis using ‘Hospitalization alert’ as the target node in order to determine which - as yet unobserved – factors have the greatest impact and hence which we should look out for next.

A major advantage of a BN model is that it can still generate predictions with incomplete information. Thus, for any nodes for which the user does not (or cannot) provide observations, the model is able to use prior probabilistic information rather than make particular assumptions.

So, although body temperature and oxygen saturation are key measurements, the user decides whether or not these measurements are actually done. Using the BN it is also possible to predict which feature will be the most informative one in contributing to the diagnosis, and this feature can be used to request additional information from the user after some initial input.

Because of the rapidly changing empirical information about all of the factors in the model the probability table parameters are being continually updated to reflect the state of knowledge. Parameters that either have no relevant empirical data, or are not logically determined, are simply estimated.

The full BN model presented in this paper is available at http://www.eecs.qmul.ac.uk/~norman/Models/covid19_for_contact_tracing_paper.ast. It can be opened and run in the free trial version of AgenaRisk (Agena Ltd 2020) from www.agenarisk.com. All the model prior and conditional probability tables can be inspected once the model is opened.

There is also an updated and extended version of the model (which can be run using the free version of AgenaRisk) incorporating many more background risk factors and symptoms (Prodhan and Fenton 2020; Butcher and Fenton 2020) is available at http://www.eecs.qmul.ac.uk/~norman/Models/covid19_for_web.cmpx

## 4. Design of the BayesCOVID Surveillance Framework

Use of our BN model provides a foundation for population surveillance of the geographical outbreak and spread of COVID19. The proposed infrastructure for personalised COVID19 status feedback and collecting geographical data is shown in Figure 5, and is inspired by related research of the authors’ research groups (van der Heijden, M. et al. 2013; Velikova, M. V. et al. 2014)

As Figure 9 illustrates, the BN is embedded or integrated into an app accessible from the user’s smartphone. This solution operates within the CardiPro environment using the Web/PWA front-end and Agena CloudAPI (S. McLachlan et al. 2020). Figure 11 shows how the user inputs their information on a smartphone and gets feedback in the form of probability, alerts and recommendations that are extracted from the underlying BN model.

**Figure 9.**
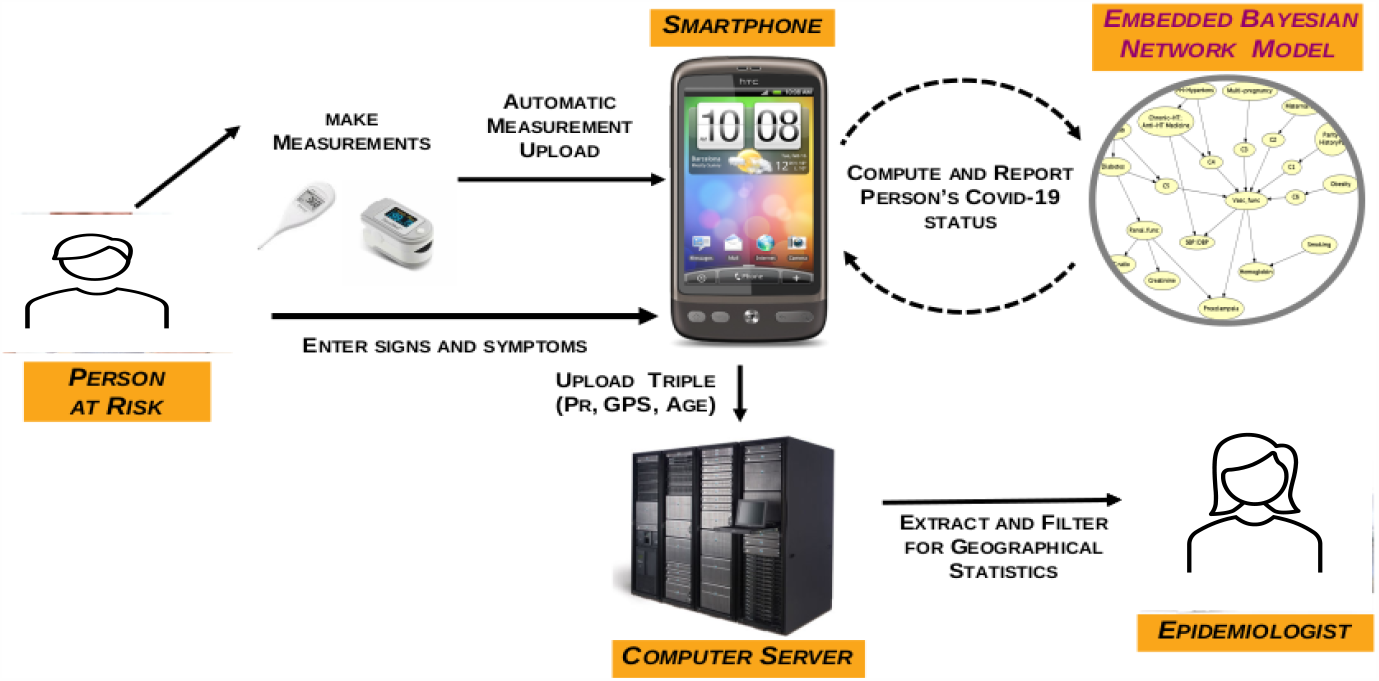
Infrastructure for personalised COVID19 feedback and collecting geographical COVID19 data

**Figure 10.**
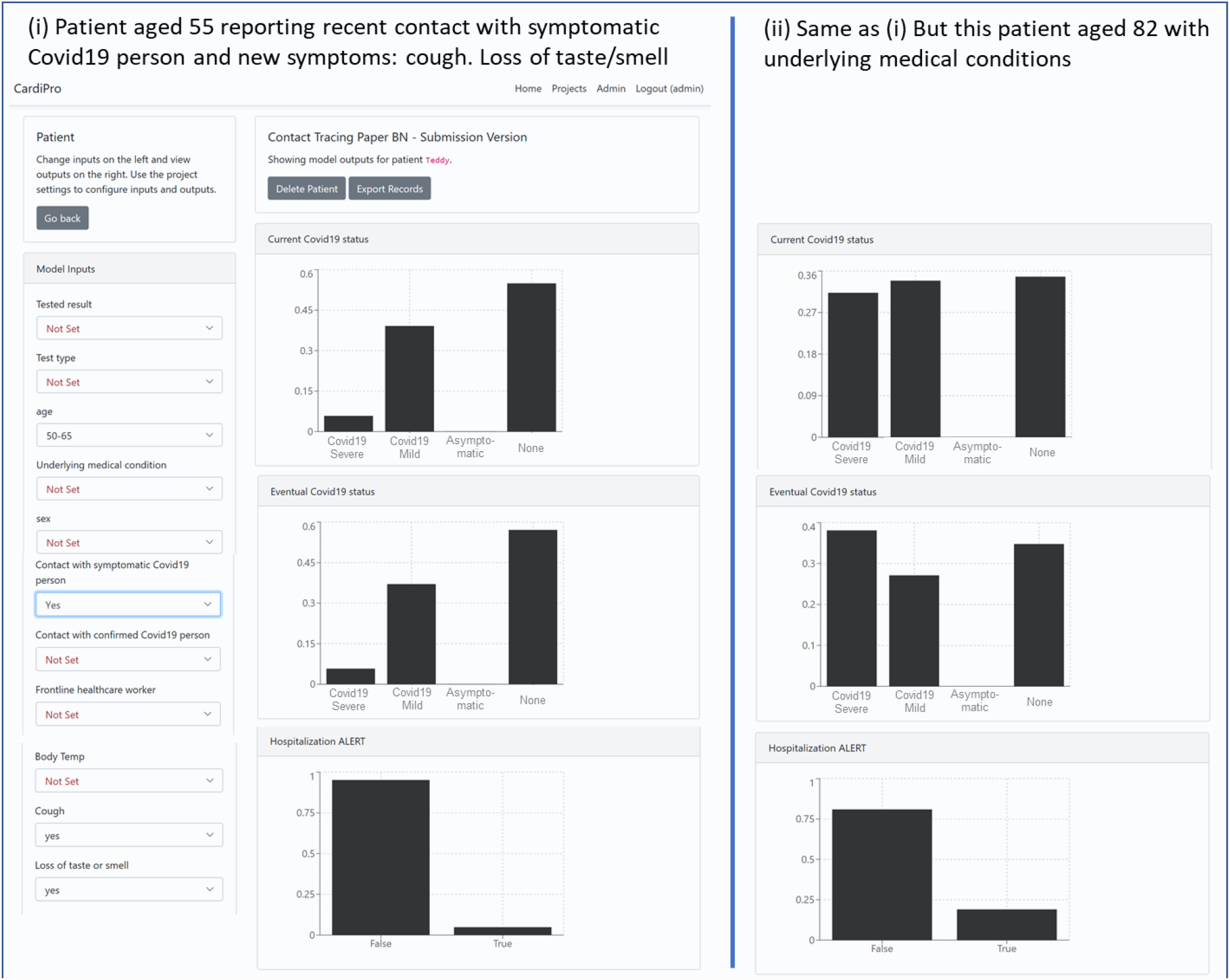
The BN model running on a mobile phone in the CardiPro Environment

Our research group has the means now to demonstrate both the elements and the entire solution presented in Figure 9^7^. The minimalist data transmitted to the server, even if coupled with collecting a similar anonymous symptom set as used for the COVID19 symptom tracker app (Drew et al. 2020), means that it is privacy preserving.

While it is assumed that a citizen of a country obtains feedback about the likelihood of the presence of asymptotic, mild or severe COVID19 from a smartphone app, the main purpose of making an app with the BN embedded is to monitor the population for detecting new outbreaks and the locations at which this occurs as early as possible. For this purpose, the only data needed to be collected centrally is the probability the user has COVID19 and their anonymised app-specific unique ID, which could also be used to follow the progress of COVID19 in the individual (possibly until hospital admission). In addition, where a person later self-identifies as positive, it might also be necessary to collect and distribute a time-stamped identifier for recent locations they have attended so that this information can be anonymously shared with other people who were present at those locations at similar times. This is how the Apple/Google APIs are intended to be used, although there is suggestion in the media that as various agencies have developed applications using these APIs, location datasets have also been made available to third parties, including advertisers, by at least three quarters of all CTA (Hautala 2020; Melendez 2020). However, collecting only the above-mentioned data triple has the advantage of minimal infringement of privacy.

## 5. Limitations of the current BN model solution^8^

Beyond the limitations already discussed in Section 3, the proposed BN model solution suffers from some limitations that are common to all BN models intended for medical decision support (including diagnosis, treatment or general risk assessment) of specific conditions. In particular:

- The risk factors, symptoms, and tests – together with the dependency relationships between them – are changing all of the time (and especially so for a new condition like COVID19) with new ones continually identified. As new empirical data about risk factors, symptoms, and tests (that are already in the model) emerge, updating the model involves only changing the relevant NPTs. As long as new symptoms do not change the dependencies in the BN (which we believe is likely in this case), the only limitation is that you cannot enter them when observed. We believe the most significant symptoms are known and have been included in the model. However, we have shown it is relatively straightforward to add new risk factors (Prodhan and Fenton 2020) and symptoms (Butcher and Fenton 2020) when they are identified; it involves adding nodes according to the existing idioms-based structure. For new symptom nodes this is simple, since they have no ‘child’ nodes and hence require only the new associated NPT based on the likelihood data for the symptom with respect to COVID19 and similar conditions. This process of adding new symptoms (which were incorporated into the extended version of the model) is described in [ref]. It is more difficult to add new risk factor nodes since doing so requires a change to the NPT for both the combined ‘risk factors’ and the ‘risk conditioned on age’ nodes; the change has to take account of the relative impact of the new risk factor with respect to existing ones. The process of adding new risk factors (which were incorporated into the extended version of the model) is described in [ref]. Adding a new type of test – and the respective test accuracy - simply involves adding a state to the node ‘Test type’ and an addition column to its child node ‘Tested result’.
- Because the ‘causal structure’ of the model is defined simply by a combination of expert judgement and common sense (albeit based on a long established idioms-based approach), it is difficult to formally justify and validate the model beyond establishing that a) its estimates reflect reality; and b) it provides better support for decision making than alternative predictive models. Inevitably, the model is a simplification and cannot address all correlations between variables and unobserved ‘errors’ (endogeneity). But the endogeneity of some specific variables – such as those associated with the testing – is explicitly handled through what is known as the measurement idiom (Kyrimi et al. 2020). Moreover, the same idiom is implicitly incorporated into the symptom nodes.

There are also further limitations that are more specific to the proposed solution. In particular:

- Where members from the same family are using the app, assumptions that users are individually and independently distributed might not be satisfied. However, the ‘infection possibility’ part of the model allows different members of the same household to input relevant information that is common to the household.
- Because most of the information required as input to the model is necessarily self-reported, there is no efficient way to check its validity without compromising user privacy. While this is true of all self-diagnosis medical BNs, it does create a potentially more serious problem in this case because ultimately the objective is not simply personalised risk assessment (whereby users would have no incentive to ‘lie’ with their inputs) but also population monitoring of virus levels. So, although users can use the model without having their ‘risk probability’ uploaded the assumption is that, by default, it will be. In this case data authentication is a potential issue since, without it, malevolent actors could use the app to distort whether or not a COVID19 outbreak is occurring in a particular location.
- There are important temporal, dynamic aspects of the problem that are only very crudely handled in the model. The model essentially incorporates just three distinct temporal phases for those who do become infected:
  - The pre-infection phase (which itself has a crude 2-phase distinction between less and more than 8 weeks for those previously infected or formally tested)
  - The period of the first 5 days of infection
  - The period after the first 5 days of infection up to the eventual outcome
  This is not a major concern because it is not assumed that users would be regularly updating their inputs. However, we recognise that some small accuracy improvements could be gained by a finer grained temporal dynamic model (e.g. with separate explicit days for each of the first 14 days of infection). However, this would require a model of much greater complexity.
- The model is inevitably highly sensitive to the assumed prior population infection rate. The version of the model presented here assumed this prior was 5% which was relevant when the virus was near its peak at the end of May 2020. However, it is simple to change this prior as it requires only a routine change to the NPT of the node ‘infected with COVID19’.

It is important to note that it was always assumed there would be regular updates to the model. Users of the model automatically access whatever is the latest version.

## 6. Conclusion

We are sceptical that any standalone CTA, manual or automated, could contain a high-prevalence highly contagious disease like COVID19. This is primarily because the CTA acts retrospectively, advising the user they were previously in close contact with an infected. In the case of COVID19, this advice often comes only after they have already begun asymptomatically shedding the disease. In contrast, the BN solution we propose integrates retrospective information with symptom tracking. It is different to other proposed solutions and models, since:

- It combines multiple different types of information into a causal model of the process that first leads to potential infection, including the possibility of immunity from previous infection, and then – based on personal risk factors - the extent to which an infected person will suffer.
- Observations of any test results and symptoms are used to update the probability of infection (and its seriousness), but the causal structure of the model also provides explanations for observations (such as why a particular test result may produce a probability of infection that seems counterintuitive).
- It provides personalised probabilities of past, current and future COVID19 status distinguishing between the probability of asymptomatic, mild or serious). The more information the user provides, the more ‘personalized’ the results – it is entirely up to the user how much information they wish to provide; the model produces probability outputs for every variable that is ‘unobserved’.
- There is minimal infringement of privacy because the personal information is not shared at all when used for personalised assessment. Moreover, even if the model is used as part of an embedded national CTA service as proposed in Section 4, the only data that needs to be shared is the current COVID19 probability status and the GPS location.

We believe that solutions like the one proposed here can have a very beneficial effect on containing the spread of this pandemic and reducing the need for draconian lockdown procedures.

## Data Availability

Links are provided in the article to data and model

http://www.eecs.qmul.ac.uk/~norman/Models/covid19_for_contact_tracing_paper.cmpx

## Acknowledgement

The authors acknowledge support from the EPSRC under project EP/P009964/1: PAMBAYESIAN: Patient Managed decision-support using Bayesian Networks and The Alan Turing Institute under the EPSRC grant EP/N510129/1. We also acknowledge the support of Agena.

## Footnotes

1 https://www.theguardian.com/world/2020/jun/18/uk-poised-to-abandon-coronavirus-app-in-favour-of-apple-and-google-models

2 https://pambayesian.org/

3 The model described in the section is NOT the latest version. The latest version can be downloaded from: http://www.eecs.qmul.ac.uk/~norman/Models/covid19_for_web.cmpx and can be run using the free trial version of AgenaRisk (www.agenarisk.com)

4 Those nodes were continuous, and their probability distributions captured the full extent of uncertainty about them. When observations about specific reported population infection and death numbers were entered, the model provided revised probability distributions for the infection and death rates as well as test accuracy.

5 As key population data changes, such as the number of people who have been tested, then we simply change the associated prior probability values in the model.

6 In AgenaRisk such ‘hidden nodes’ can literally be hidden from view.

7 A new and updated version of the web/phone app which enables the latest version of the model to be run through a simple questionnaire interface can be accessed at https://covid19.apps.agenarisk.com

8 Some of the limitations of the BN model described in this paper have been addressed in the latest version which can be downloaded from: http://www.eecs.qmul.ac.uk/~norman/Models/covid19_for_web.cmpx and run using the free trial version of AgenaRisk (www.agenarisk.com) The latest version of the model can also be run through a simple questionnaire interface over the web or on a mobile phone at https://covid19.apps.agenarisk.com/

## References

Agena Ltd. 2020. “AgenaRisk.” http://www.agenarisk.com.

Ai, Tao, Zhenlu Yang, Hongyan Hou, Chenao Zhan, Chong Chen, Wenzhi Lv, Qian Tao, Ziyong Sun, and Liming Xia. 2020. “Correlation of Chest CT and RT-PCR Testing in Coronavirus Disease 2019 (COVID-19) in China: A Report of 1014 Cases.” Radiology, February, 200642. https://doi.org/10.1148/radiol.2020200642.

Apea, Vanessa J, Yize I Wan, Rageshri Dhairyawan, Zudin A Puthucheary, Rupert M Pearse, Chloe M Orkin, and John R Prowle. 2020. “Ethnicity and Outcomes in Patients Hospitalised with COVID-19 Infection in East London: An Observational Cohort Study.” MedRxiv, June, 2020.06.10.20127621. https://doi.org/10.1101/2020.06.10.20127621.

Beeching, Nick J, Tom E Fletcher, and Mike B J Beadsworth. 2020. “Covid-19: Testing Times.” BMJ (Clinical Research Ed.) 369 (April): m1403. https://doi.org/10.1136/bmj.m1403.

Bengali, S. 2020. “He Was Symptom-Free. But the Coronavirus Stayed in His Body for 40 Days.” LA Times, May 4, 2020. https://www.latimes.com/world-nation/story/2020-04-30/why-some-patients-keep-testing-positive-for-the-coronavirus.

Butcher, Rachel, and Norman E. Fenton. 2020. “Extending the Range of Symptoms in a Bayesian Network for the Predictive Diagnosis of COVID-19.” https://doi.org/links_side_Button.jpg (3757 bytes) default_top_Bar.jpg (16198 bytes) Books Patents Papers Books Fenton, N.E. and M. Neil, Risk Assessment and Decision Analysis with Bayesian Networks, Second Edition. 2018, Chapman and Hall/CRC Press, ISBN: 9781138035119, 2018

Fenton, N. E. and J. Bieman (2014) “Software Metrics: A Rigorous and Practical Approach” (3rd Edition). CRC Press, ISBN 9781439838228 Fenton, N.E.and M. Neil, Risk Assessment and Decision Analysis with Bayesian Networks. 2012, CRC Press, I.

Carver, C., and N. Jones. 2020. “Comparative Accuracy of Oropharyngeal and Nasopharyngeal Swabs for Diagnosis of COVID-19.” https://www.cebm.net/covid-19/comparative-accuracy-of-oropharyngeal-and-nasopharyngeal-swabs-for-diagnosis-of-covid-19/.

Centers for Disease Control and Prevention. 2020. “Symptoms of Coronavirus | CDC.” 2020. https://www.cdc.gov/coronavirus/2019-ncov/symptoms-testing/symptoms.html.

Chua, Felix, Darius Armstrong-James, Sujal R Desai, Joseph Barnett, Vasileios Kouranos, Onn Min Kon, Ricardo José, et al. 2020. “The Role of CT in Case Ascertainment and Management of COVID-19 Pneumonia in the UK: Insights from High-Incidence Regions.” The Lancet. Respiratory Medicine 8 (5): 438–40. https://doi.org/10.1016/S2213-2600(20)30132-6.

Collaborative, The OpenSAFELY, Elizabeth Williamson, Alex J Walker, Krishnan J Bhaskaran, Seb Bacon, Chris Bates, Caroline E Morton, et al. 2020. “OpenSAFELY: Factors Associated with COVID-19-Related Hospital Death in the Linked Electronic Health Records of 17 Million Adult NHS Patients.” MedRxiv, May, 2020.05.06.20092999. https://doi.org/10.1101/2020.05.06.20092999.

Constantinou, Anthony Costa, Norman E Fenton, William Marsh, and Lukasz Radlinski. 2016. “From Complex Questionnaire and Interviewing Data to Intelligent Bayesian Network Models for Medical Decision Support.” Artificial Intelligence in Medicine 67 (January): 75–93. https://doi.org/10.1016/j.artmed.2016.01.002.

Cowell, RG, AP Dawid, SL Lauritzen, and DJ Spiegelhalter. 1999. Probabilistic Networks and Expert Systems. New York: Springer.

Day, Michael. 2020. “Covid-19: Identifying and Isolating Asymptomatic People Helped Eliminate Virus in Italian Village.” BMJ (Clinical Research Ed.) 368 (March): m1165. https://doi.org/10.1136/bmj.m1165.

Drew, David A, Long H Nguyen, Claire J Steves, Cristina Menni, Maxim Freydin, Thomas Varsavsky, Carole H Sudre, et al. 2020. “Rapid Implementation of Mobile Technology for Real-Time Epidemiology of COVID-19.” Science (New York, N.Y.) 368 (6497): 1362–67. https://doi.org/10.1126/science.abc0473.

Fenton, Norman E., Martin Neil, Magda Osman, and Scott McLachlan. 2020. “COVID-19 Infection and Death Rates: The Need to Incorporate Causal Explanations for the Data and Avoid Bias in Testing.” Journal of Risk Research, April, 1–4. https://doi.org/10.1080/13669877.2020.1756381.

Fenton, Norman E. 2020. “A Note on ‘Collider Bias Undermines Our Understanding of COVID-19 Disease Risk and Severity’ and How Causal Bayesian Networks Both Expose and Resolve the Problem,” May. http://arxiv.org/abs/2005.08608.

Fenton, Norman E, and Martin Neil. 2018. Risk Assessment and Decision Analysis with Bayesian Networks. 2nd ed. CRC Press, Boca Raton.

Griffith, Gareth, Tim T Morris, Matt Tudball, Annie Herbert, Giulia Mancano, Lindsey Pike, Gemma C Sharp, et al. 2020. “Collider Bias Undermines Our Understanding of COVID-19 Disease Risk and Severity.” MedRxiv, May, 2020.05.04.20090506. https://doi.org/10.1101/2020.05.04.20090506.

Hautala, L. 2020. “COVID-19 Contact Tracing Apps Create Privacy Pitfalls around the World.” CNET. 2020. https://www.cnet.com/news/covid-contact-tracing-apps-bring-privacy-pitfalls-around-the-world/

Heijden, M., van der, P. J. Lucas, B. Lijnse, Y. F. Heijdra, and T. R. Schermer. 2013. “An Autonomous Mobile System for the Management of COPD.” Journal of Biomedical Informatics 46 (3): 458–69.

Heneghan, Carl, Jon Brassey, and Tom Jefferson. 2020. “COVID-19: What Proportion Are Asymptomatic?” Oxford. https://www.cebm.net/covid-19/covid-19-what-proportion-are-asymptomatic/.

Kyrimi, E., M. Neves, M. Neil, W. Marsh, S. McLachlan, and Norman E Fenton. 2020. “Medical Idioms for Clinical Bayesian Network Development.” Artificial Intelligence in Medicine 108 (103495). https://doi.org/10.1016/j.jbi.2020.103495.

Long, Quan-Xin, Xiao-Jun Tang, Qiu-Lin Shi, Qin Li, Hai-Jun Deng, Jun Yuan, Jie-Li Hu, et al. 2020. “Clinical and Immunological Assessment of Asymptomatic SARS-CoV-2 Infections.” Nature Medicine, June, 1–5. https://doi.org/10.1038/s41591-020-0965-6.

McLachlan, S., H. Paterson, K. Dube, E. Kyrimi, E. Dementiev, M. Neil, B. Daley, G.A. Hitman, and N. Fenton. 2020. “Real-Time Online Probabilistic Medical Computation Using Bayesian Networks.” https://easychair.org/publications/preprint/9Jks.

McLachlan, Scott, Peter Lucas, Kudakwashe Dube, Graham A Hitman, Magda Osman, Evangelia Kyrimi, Martin Neil, and Norman E Fenton. 2020. “Bluetooth Smartphone Apps: Are They the Most Private and Effective Solution for COVID-19 Contact Tracing?,” May. http://arxiv.org/abs/2005.06621.

McLachlan, Scott, Peter Lucas, Kudakwashe Dube, Graham Scott McLachlan, Graham A Hitman, Magda Osman, Evangelia Kyrimi, Martin Neil, and Norman E Fenton. 2020. “The Fundamental Limitations of COVID-19 Contact Tracing Methods and How to Resolve Them with a Bayesian Network Approach.” London, UK. https://doi.org/10.13140/RG.2.2.27042.66243.

Melendez, S. 2020. “North Dakota’s COVID-19 App Has Been Sending Data to Foursquare and Google.” Fastcompany. 2020. https://www.fastcompany.com/90508044/north-dakotas-covid-19-app-has-been-sending-data-to-foursquare-and-google.

Michelen, Melina, Nicholas Jones, and Charitini Stavropoulou. 2020. “In Patients of COVID-19, What Are the Symptoms and Clinical Features of Mild and Moderate Cases?” Oxford. https://www.cebm.net/covid-19/in-patients-of-covid-19-what-are-the-symptoms-and-clinical-features-of-mild-and-moderate-case/.

Mizumoto, Kenji, Katsushi Kagaya, Alexander Zarebski, and Gerardo Chowell. 2020. “Estimating the Asymptomatic Proportion of Coronavirus Disease 2019 (COVID-19) Cases on Board the Diamond Princess Cruise Ship,Yokohama, Japan, 2020.” Eurosurveillance 25 (10): 2000180. https://doi.org/10.2807/1560-7917.ES.2020.25.10.2000180.

Neil, Martin, Norman E Fenton, and Lars Nielson. 2000. “Building Large-Scale Bayesian Networks.” The Knowledge Engineering Review 15 (3): 257–84. https://doi.org/10.1017/S0269888900003039.

Neil, Martin, Norman E Fenton, Magda Osman, and Scott McLachlan. 2020. “Bayesian Network Analysis of Covid-19 Data Reveals Higher Infection Prevalence Rates and Lower Fatality Rates than Widely Reported.” Journal of Risk Research, May. https://doi.org/10.1080/13669877.2020.1778771.

Osman, M, Fenton, S McLachlan, P. Lucas, K. Dube, G. A. Hitman, E Kyrimi, and M Neil. 2020. “The Thorny Problems of Covid-19 Contact Tracing Apps: The Need for a Holistic Approach.” Journal of Behavioral Economics for Policy 4: 43–59.

Pearl, Judea. 1988. Probabilistic Reasoning in Intelligent Systems: Networks of Plausible Inference. San Francisco: Morgan Kaufmann Publishers Inc.

Prodhan, Georgina, and Norman E. Fenton. 2020. “Extending the Range of COVID-19 Risk Factors in a Bayesian Network Model for Personalised Risk Assessment.” https://doi.org/10.1101/2020.10.20.20215814.

Servick, Kelly. 2020. “COVID-19 Contact Tracing Apps Are Coming to a Phone near You. How Will We Know Whether They Work?” Science, May. https://doi.org/10.1126/science.abc9379.

Velikova, M. V., J. A. Terwisscha van Scheltinga, P. J. Lucas, and M. Spaanderman. 2014. “Exploiting Causal Functional Relationships in Bayesian Network Modelling for Personalised Healthcare.” International Journal of Approximate Reasoning 55: 59–73.

Yang, Yang, Minghui Yang, Chenguang Shen, Fuxiang Wang, Jing Yuan, Jinxiu Li, Mingxia Zhang, et al. 2020. “Evaluating the Accuracy of Different Respiratory Specimens in the Laboratory Diagnosis and Monitoring the Viral Shedding of 2019-NCoV Infections.” MedRxiv, February, 2020.02.11.20021493. https://doi.org/10.1101/2020.02.11.20021493.

